# DRUG-RESISTANT TUBERCULOSIS CASE-FINDING STRATEGIES: A SCOPING REVIEW PROTOCOL

**DOI:** 10.1101/2022.05.30.22275779

**Authors:** Susanna S van Wyk, Marriott Nliwasa, James A Seddon, Graeme Hoddinott, Lario Viljoen, Emmanuel Nepolo, Gunar Günther, Nunurai Ruswa, Hsien-Ho Lin, Stefan Niemann, Neel Gandhi, Sarita Shah, Mareli Claassens

**Affiliations:** Centre for Evidence Based Health Care, Division of Epidemiology and Biostatistics, Department of Global Health, Stellenbosch University, Cape Town, South Africa; Helse Nord Tuberculosis Initiative, Kamuzu University of Health Sciences, Blantyre, Malawi; Desmond Tutu TB Centre, Department of Paediatrics and Child Health, Faculty of Medicine and Health Sciences, Stellenbosch University, Cape Town, South Africa; Department of Infectious Diseases, Imperial College London, London, United Kingdom; Department of Human, Biological and Translational Medical Science, School of Medicine, University of Namibia, Windhoek, Namibia; Department of Pulmonary Medicine, Inselspital, Bern University Hospital, University of Bern, Bern, Switzerland; Department of Human, Biological & Translational Medical Science, School of Medicine, University of Namibia, Windhoek, Namibia; Clinical and Programmatic Coordinator for Drug Resistant Tuberculosis, Ministry of Health and Social Services, Windhoek, Namibia; Institute of Epidemiology and Preventive Medicine, National Taiwan University, Taiwan; Molecular and Experimental Mycobacteriology Group and National Reference Center for Mycobacteria, Forschungszentrum Borstel, Germany; Departments of Epidemiology and Global Health, Rollins School of Public Health, Emory University, Atlanta, USA

**Keywords:** Tuberculosis, drug-resistant tuberculosis, drug-resistant tuberculosis case finding, drug-resistant tuberculosis case detection, drug-resistant tuberculosis screening

## Abstract

**Background:** Transmission of drug-resistant tuberculosis (TB) is ongoing, and many households face catastrophic costs when a member is diagnosed and treated for drug-resistant TB. Finding individuals with drug-resistant TB and initiating treatment as early as possible is important to improve patient clinical outcomes and to break the chain of transmission to help control the pandemic.

To our knowledge systematic reviews assessing effectiveness, cost-effectiveness, acceptability and feasibility of different drug-resistant TB case-finding strategies to inform research, policy and practice, have not been conducted and it is unknown whether enough research exists to conduct such reviews. It is also unknown whether case-finding strategies are similar for drug-resistant TB and drug-susceptible TB and whether we can draw on findings from drug-susceptible reviews to inform decisions on drug-resistant TB case-finding strategies.

**Methods:** The question for our review is: what literature is available on drug-resistant TB case finding and which case-finding strategies are described? We will look at studies that have sought to improve drug-resistant TB case detection. We will search the academic databases of CENTRAL, MEDLINE and EMBASE using no language or date restrictions. We will apply broad criteria for screening titles, abstracts and full text articles. A data extraction form will be developed in excel and applied to all primary research reports to collect standard information on each study. We will provide a narrative report with supporting figures and/or tables to summarize the data. A systems-based logic model, developed from a synthesis of drug-susceptible TB case-finding strategies, will be used as a framework to describe different strategies, resulting pathways and enhancements of pathways.

**Discussion:** This scoping review will chart existing literature on drug-resistant TB case finding and identify priority question(s) for a systematic review. It will also describe drug-resistant TB case-finding strategies and how they fit into a model of drug-susceptible TB case-finding pathways. The review will guide further research to inform decisions on drug-resistant TB case finding policy and practice.

## BACKGROUND

With the emergence of *Mycobaterium tuberculosis (M. tb)* strains resistant to first line anti-tuberculosis drugs, strategies to control tuberculosis (TB) became even more challenging.^1^ It is estimated that almost half a million people developed rifampicin-resistant (RR) TB, of which 78% had multidrug-resistant TB (MDR-TB) in 2019.^2^ Although drug-resistant TB is not as prevalent as drug-susceptible TB, it is more difficult to diagnose, treatment is longer and more toxic, outcomes are worse and costs are higher. Sixty-seven to 100% of people with drug-resistant TB in their households face catastrophic costs (total costs equivalent to >20% of annual household income).^2^

Finding individuals with drug-resistant TB and initiating treatment as early as possible is important to improve patient clinical outcomes and to break the chain of transmission to help control the pandemic. But despite new diagnostic technologies, only 38% of the estimated number of people who developed drug-resistant TB initiated treatment in 2019.^2,3^ TB can be detected following the patient presenting passively to health services or following one of several different screening pathways depending on the case-finding strategy of a TB programme.^4^ Pathways can also be enhanced via several activities such as health promotion in the community, improved access to TB diagnostic services or training of health workers to identify presumptive TB at general health services. Multiple activities often result in complex interventions and heterogeneous trials which are difficult to meta-analyse in systematic reviews.^5,6^

We will therefore conduct a scoping review to chart existing literature on drug-resistant TB case finding and identify priority question(s) for a systematic review.^7,8^ We will also describe existing strategies and how they fit into a model of drug-susceptible TB case-finding pathways.

## METHODS

The Arksey and O’Malley framework^9,10^, the Joanna Briggs Institute scoping review methodology^8^ and the PRISMA extension for scoping reviews (PRISMA-ScR)^11^ will guide methods for this scoping review.

### Defining the research question

The question for our review is: what literature is available on drug-resistant TB case finding and which case-finding strategies are described? We will look at studies that have sought to improve drug-resistant TB case detection.

### Identifying relevant studies

We will use the following eligibility criteria:

**Inclusion criteria**

- **Participants** Participants can be any age or gender. They can be people receiving an intervention or providers of an intervention.
- **Concept** Intervention strategies aiming to improve or enhance participants’ pathway(s) to drug-resistant TB case detection specifically. Intervention strategies aiming to improve TB case finding in general and only report on the yield of drug-resistant TB cases will be excluded.
- **Context** Studies conducted at primary care, secondary care, tertiary care or community settings. We will exclude studies that are purely laboratory based, but will include studies where screening of sputum samples is part of an intervention to improve drug-resistant TB case finding.
- **Study designs** Systematic reviews, excluding narrative reviews. Primary studies irrespective of study design. Qualitative studies, where the experiences of individuals who receive the intervention or those who provide the intervention are investigated. We will include studies of diagnostic test accuracy if the study describes a drug-resistant TB screening strategy and we will include trials comparing different screening or diagnostic tools within case-finding interventions. We will exclude prevalence surveys, except if the survey includes an intervention strategy to find drug-resistant TB cases specifically. We will exclude editorials, opinion articles, meeting summaries and guidelines.

We will search the academic databases of Medline (PubMed), Embase (Ovid), The Cochrane Library, Africa-Wide Information (EBSCOhost), CINAHL (EBSCOhost), Epistemonikos and PROSPERO using no language or date restrictions. Reference lists of included studies will be searched.

The preliminary search string will include combinations of the following three domains:

- terms related to “tuberculosis”
- terms related to “drug resistance”
- terms related to “case finding”, “case detection”, “screening”, “contact investigation” and “contact tracing”

Appropriate MesH terms will be added to the different databases. The search strategy will be piloted and refined in consultation with an information specialist. We will also contact experts working in the field to collect information about ongoing primary research or relevant research missed by the electronic search.

### Study selection

We will use Rayyan systematic review software ^12^ to screen titles, abstracts and full text articles. Four reviewers (SvW, MN, LV and MC) will screen abstracts in duplicate for inclusion. They will resolve conflicts via discussion and meet at the beginning and midpoint of abstract screening to discuss challenges and possible refinement of the search strategy. Three reviewers (SvW, LV and MN) will then screen full text articles for inclusion. Disagreements will be resolved with a third reviewer (MC) to determine final inclusion.

### Charting the data

We will develop a data extraction form in excel. The data extraction form will be applied to all primary research reports to collect standard information on each study. Information will include:

- Authors, journal, year of publication
- Aim/purpose of the research
- Study design
- Country: income, TB prevalence, human immunodeficiency virus (HIV) prevalence; urban/rural setting
- Participants: age, sex, HIV status, other reported risk factors
- Target group and how the group was identified if applicable
- Intervention(s): All components (activities) of the intervention, type(s) of providers and screening and diagnostic tools used
- Treatment support, including preventive therapy
- Outcomes assessed

Five authors (SvW, MN, LV, MC and GH) will extract data, one author per paper. A second reviewer (SvW/MC) will check extracted data from each study. The data extraction team and other co-authors will meet regularly after each 5-10 studies to determine whether their approach is consistent and in line with the research question.

### Collating, summarizing and reporting the results

We will provide a narrative report with supporting figures and/or tables to summarize the data. Table 1 contains definitions we will use in charting, collating, summarizing and reporting our results.

**Table 1:** Definitions.

|  |  |
| --- | --- |
| **Drug-resistant TB** | All types of drug-resistant TB: single drug-resistant TB, multidrug-resistant TB, extensively drug-resistant TB and any other drug-resistant TB reported by the authors. |
| **Systematic screening for active TB** | "The systematic identification of people with suspected (presumptive) active TB, in a predetermined target group, using tests, examinations or other procedures that can be applied rapidly. Among those screened positive, the diagnosis needs to be established by one or several diagnostic tests and additional clinical assessments, which together have high accuracy."<sup>3</sup> |
| **A screening tool** | Tests, examinations or other procedures used for systematic screening for active TB. Examples of TB screening tools include a structured symptom-based questionnaire, chest radiography (CXR), or an algorithm.<sup>4</sup> Algorithms may include sequential or parallel tests. With sequential tests, only those who screen positive with the initial test receive a second test. With parallel tests, those who screen positive on any of the tests are regarded as screen positives. |
| **A diagnostic tool** | Tests, examinations or other procedures used to establish a diagnosis of TB in people identified with presumptive TB. Examples of TB diagnostic tools include a clinical algorithm, sputum smear microscopy, Xpert MTB/RIF or culture.<sup>4</sup> |
| **TB symptom(s)** | Any TB symptom, e.g. cough, fever, night sweats, weight loss or combination of TB symptoms as defined by the study authors. |
| **Care seeking** | People seeking care for a perceived health problem. |
| **TB care seeking** | People seeking care for TB symptoms specifically. |
| **A risk group** | Any group of people in whom the prevalence or incidence of TB is significantly higher than in the general population. Examples of risk groups include a whole population within a geographical area or TB contacts.<sup>3</sup> |
| **A clinical risk group** | Individuals diagnosed with a specific disease or condition that increases their risk for TB, e.g. people living with HIV (PLHIV). |
| **Presumptive TB** | Presumptive TB is identified when a provider identifies a patient with suspected active tuberculosis. In the context of screening, a person who screens positive is a presumptive TB case. |
| **Passive case finding**<br>**Passive case finding with an element of systematic screening**<br>**Triage**<br>**Enhanced case finding**<br>**Active case finding**<br>**Contact tracing/ contact investigation**<br>**Intensified case finding** | *Some of these definitions overlap and are used inconsistently within the literature. We will therefore use our logic model (Fig 1) to clearly describe pathways rather than labelling them with these terms. |
| * Van Wyk SS, Medley N, Young T and Oliver S. Repairing boundaries along pathways to tuberculosis case detection: An evidence synthesis. Work submitted for publication, June 2021. |  |

A systems-based logic model developed from a synthesis of drug-susceptible TB case-finding strategies (Figure 1) will be used as a framework to describe different strategies and resulting pathways. Enhancements of pathways will also be described and may include enhanced care seeking (e.g. health promotion), improved access to care for those seeking care (e.g. mobile clinics), improved access to TB screening (e.g. incentives), improved identification of presumptive TB by health workers (e.g. training of health workers, incentives) and improved access to TB diagnostic services for individuals identified with presumptive TB (e.g. transport, sputum collection in the community, mobile laboratory). For screening pathways we will report on target groups and screening and diagnostic tools used. Quality appraisal will not be conducted, because this is a scoping review and our interest is in the existing evidence base, regardless of study design and quality.

**Figure 1:**
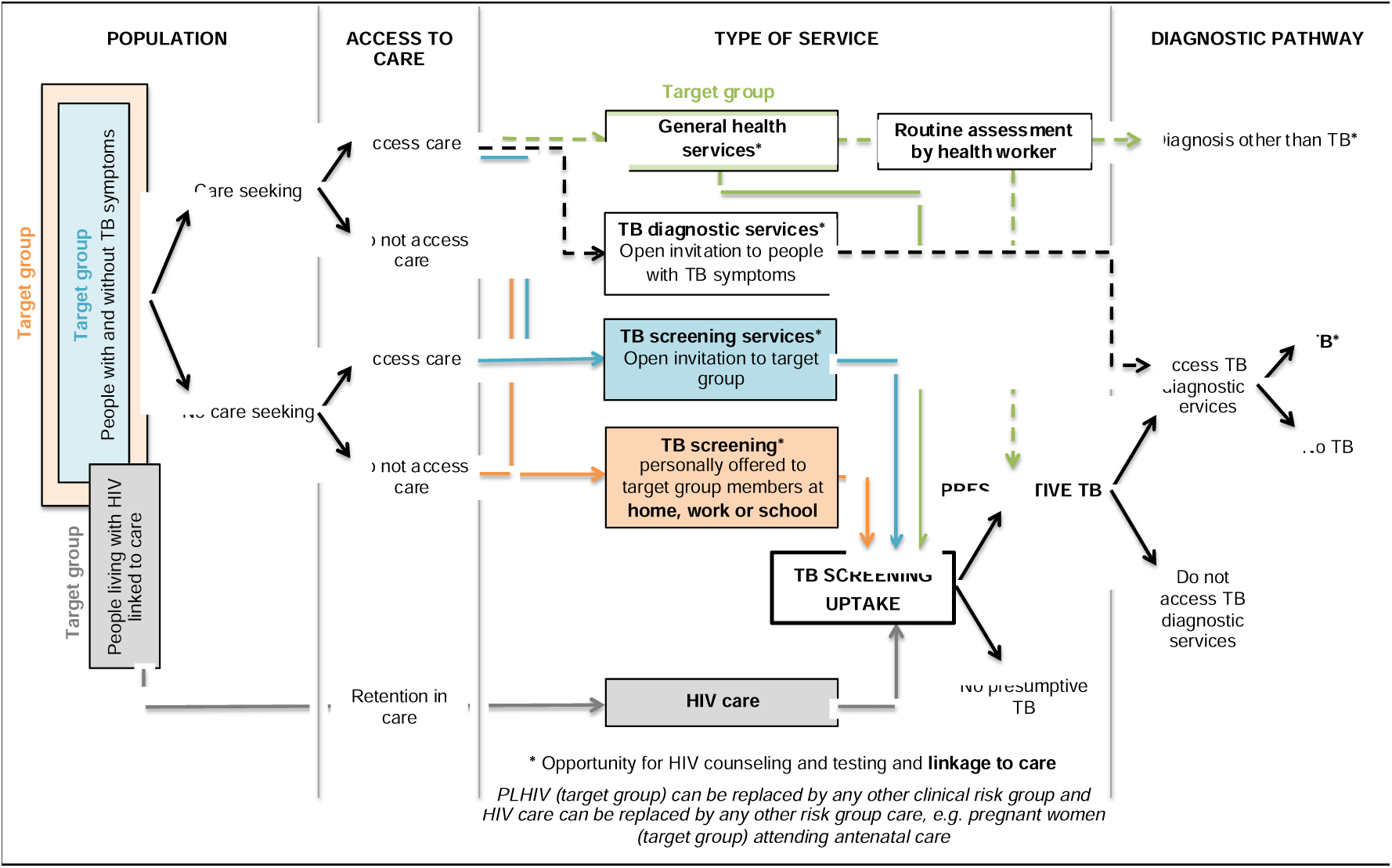
A systems-based logic model depicting types of services and associated pathways to TB case detection. The model distinguishes six pathways to TB case detection, namely two care-seeking pathways (green and black dashed lines) and four screening pathways (green, blue, orange and grey solid lines). People perceiving themselves to have a health problem and access general health services, follow the general care-seeking pathway where a provider can identify presumptive TB on routine assessment, i.e., history taking and clinical examination, of an individual patient (green dashed line). People perceiving themselves to have TB symptoms may also follow the specific TB care-seeking pathway to TB diagnostic services, where all people accessing care are evaluated for possible active TB (black dashed line). People invited to TB services regardless of symptoms follow TB screening pathways and may be identified with presumptive TB even if they do not seek care for TB symptoms. Four screening pathways are distinguished: TB screening offered to all people accessing general health services (green solid line), dedicated TB screening services at a health facility or mobile clinic with open invitation to a whole population or TB contacts (blue solid line), TB screening offered to target group members at home, work or school (orange solid line) and TB screening offered to people living with HIV linked to care (grey solid line). A person who screens positive on the TB screening pathway is identified as a presumptive TB case and should receive confirmation of a diagnosis by accessing TB diagnostic services.

## DISCUSSION

This scoping review will chart existing literature on drug-resistant TB case finding and identify priority question(s) for a systematic review. It will also describe drug-resistant TB case-finding strategies and how they fit into a model of drug-susceptible TB case-finding pathways.

### Strengths and limitations

Our multidisciplinary review team consists of researchers with extensive experience in TB related research and the conduct of systematic reviews and qualitative evidence synthesis. Their experience would be invaluable in collating and summarizing diverse literature in a sensible way. Another strength is the use of a systems-based logic model that was developed from a synthesis of drug-susceptible TB case-finding strategies. The model will help us to construct meaningful pathway descriptions for possible comparisons in future research and to assess whether drug-resistant TB case-finding pathways are similar to drug-susceptible pathways.

One limitation to the review is the diverse and inconsistent use of intervention terminology within the literature, which may result in missing relevant studies. Although we cannot overcome this problem in full, we will work with an information specialist to pilot and optimize our search strategy and we will discuss potential refinement of our search at regular meetings during the screening phase. Poor reporting of intervention strategies may also cause misunderstanding and misclassification of interventions. However, we do not assess effectiveness as an outcome and therefore bias due to misclassification will be a minor issue. Lastly, drug-resistant TB case-finding strategies may not fit into a model developed from drug-susceptible strategies. Nevertheless, such a situation will provide an opportunity to refine the model for future research.

### Conclusion

This scoping review will chart the existing body of literature on drug-resistant TB case-finding strategies and will guide further research to inform decisions on drug-resistant TB case finding policy and practice.

## Supporting information

Prisma-P Checklist

## Data Availability

This is a protocol and data have not yet been finalised.

## LIST OF ABBREVIATIONS

TB: tuberculosis
*M. tb*: *Mycobaterium tuberculosis (M. tb)*
RR: rifampicin-resistant
MDR-TB: multidrug-resistant tuberculosis
PRISMA-ScR: PRISMA extension for scoping reviews
HIV: human immunodeficiency virus
PLHIV: people living with human immunodeficiency virus

## DECLARATIONS

### Ethics approval and consent to participate

Not applicable

### Consent for publication

Not applicable

### Availability of data and materials

Not applicable

## Authors’ contributions

MC and SvW conceived the study idea. SvW drafted the protocol. All authors read and approved the final protocol.

## Acknowledgements

Not applicable

## Additional file 1

The PRISMA-P Checklist

